# Implementation of Evidence-Based Medicine in Primary Care Through the Use of Encounter Shared Decision Making Tools: The ShareEBM Pragmatic Trial

**DOI:** 10.1101/2023.12.19.23300235

**Authors:** Annie LeBlanc, Megan E Branda, Jason Egginton, Jonathan W Inselman, Sara Dick, Janet Schuerman, Jill Kemper, Nilay D Shah, Victor M Montori

## Abstract

**BACKGROUND:** While decision aids have been proven effective to facilitate patient-centered discussion about evidence-based health information in practice and enable shared decision making (SDM), a chasm remains between the promise and the use of these SDM tools in practice.

**AIMS:** To promote evidence-based patient-centered care in primary care by using encounter SDM tools for medication management of chronic conditions.

**METHODS:** We conducted a mixed methods study centered around a practice-based, multi-centered pragmatic randomized trial comparing active implementation (active) to passive dissemination (passive) of a web-based toolkit, ShareEBM, to facilitate the uptake in primary care of four SDM tools designed for use during clinical encounters. These tools supported collaborative decisions about medications for chronic conditions. ShareEBM included activities and tactics to increase the likelihood that encounter SDM tools will be routinized in practice. Study team members worked closely with practices in the active arm to actively integrate and promote the use of SDM tools; passive arm practices received no support from the study team. The embedded qualitative evaluation included clinician phone interviews (n=10) and site observations (n=5) for active practices, and exit focus groups for all practices (n=11).

**RESULTS:** Eleven practices and 62 clinicians participated in the study. Clinicians in the active arm used SDM tools in 621 encounters (Mean [SD]: 21 [25] encounters per clinician, range: 0-93) compared to 680 in the passive arm (Mean [SD]: 20 [40] encounters per clinician, range: 0-156, p=0.4). Six of 29 (21%) clinicians in the active arm and 14 of 33 (42%) in the passive arm did not use any tools (p=0.1). Clinicians’ views covered four major themes: general views of using encounter SDM tools, perceived impact on patients, strategies used, and how encounter SDM tools are incorporated into practice flow.

**CONCLUSION:** Neither active nor passive implementation of a toolkit improved the uptake and use of encounter SDM tools in primary care. Overcoming clinician reluctance to consider using encounter SDM tools, their seamless integration into the electronic and practice workflows, and ongoing feedback about the quality of their use during encounters appear necessary to implement their use in primary care practices.

## INTRODUCTION

Shared decision making (SDM) describes the collaboration of patients and clinicians to address the problematic situation of the patient.^1^ This process benefits from the participation and engagement of both patient and clinician, the mobilization of their expertise and experience, and drawing from the best available research evidence about the available options for care. SDM tools for use during the clinical encounter can be designed and implemented to support this collaborative conversation in practice.^2,3^ These tools often present evidence from comparative effectiveness research describing the relative benefits, harms, burdens, and costs of the possible paths forward. Their effectiveness in promoting SDM has been repeatedly established in more than a hundred randomized controlled trials.^2,4^ Many such tools are available but their uptake and routine use in practice remains rare. This may reduce the quality of patient-centered care and represents an opportunity to improve care.

SDM is particularly important in the care of patients with chronic conditions. Chronic conditions affect six out of every ten Americans, significantly impact quality of life, and are the leading causes of death and disabilities.^5^ Most common chronic conditions (e.g., heart disease, diabetes, depression, osteoporosis) are managed in primary care, yet international studies report major gaps in the quality of evidence-based, patient-centered care for patients with chronic conditions.^6,7^ In patients with chronic conditions, treatments and life have ongoing interactions such that achieving a good fit is necessary. Thus, the simple prescription of a recommended therapy for each of the patient’s conditions may not produce the expected results because it may not be initiated or used consistently, or its use may end up being undesirable or unfeasible. This may lead to inappropriate, unnecessary, or unwanted care and suboptimal outcomes.^8,9^

At the time of this study, investigators at the Knowledge and Evaluation Research (KER) Unit at Mayo Clinic had over a decade of experience partnering with patients, clinicians and other stakeholders to create encounter SDM tools, decision aids designed to support collaborative conversations at the point of care.^9–15^ These decision aids have been evaluated in practical randomized trials and proven effective in supporting greater patient and clinician participation in SDM, improving patient and clinician satisfaction with care and decisions, without demanding more clinical time.^9–18^

Little is known about how to effectively embed SDM encounter tools within routine practices.^19,20,21^ Indeed, the use of decision aids has not been successfully implemented and routinized in clinical practices despite ethical desirability and growing clinical and policy consensus about its importance. Implementation of SDM and decision aids in practice has proven difficult and far more complex than merely making them available to patients and clinicians.^19,20^ This calls for implementation science to overcome the barriers that preclude their routine use and limit their desirability and widespread adoption.^21^

Here we report on the results of a practice-based study seeking to compare two implementation approaches to increase the uptake and use of effective SDM encounter tools in primary care practices.

## METHODS

### Study Design

We conducted a mixed-methods study centered around a pragmatic, practice-based, multi-centered randomized trial and an associated qualitative assessment that compared active implementation to passive dissemination of ShareEBM, an implementation toolkit to facilitate uptake of encounter SDM tools among primary care practices serving patients with chronic conditions. The Mayo Clinic Institutional Review Board approved study procedures and the trial was registered at Clinicaltrials.gov (NCT02136199). We adhered to the CONSORT and COREQ guidelines in reporting our findings.^22,23^

### Setting, Participants & Recruitment

We conducted the study in 12 rural, suburban, and inner-city primary care practices across Minnesota and Wisconsin, USA. Practices were eligible for the study if they had at least 2 primary care clinicians (defined as any healthcare professional involved in direct patient care) committed to participate in the study, a designated site champion, and endorsement from practice leadership. Practices with an established relationship with the Institute for Clinical System Improvements (ICSI), a statewide Minnesotan institution that supports health care delivery systems in quality improvement were invited to participate in this study.

Clinicians were eligible to participate in the study if they participated in the care of patients with chronic conditions and provided informed written consent to participate in this study. They were approached to participate in the study during an initial on-site meeting with members of the investigation team, and then at any point during the trial period either by a member of the research team or by the site champion.

### Allocation procedures

We randomly allocated practices to the intervention (active implementation) or control (passive dissemination) arm after they were stratified by setting (rural vs. urban) and number of clinicians (<10 vs. ≥10). The study statistician performed the random allocation centrally after practices were enrolled to ensure concealment of allocation. Investigators, practices, and clinicians were not masked to the allocation. To prevent loss to follow-up, ensure complete data collection, and support the intention-to-treat principle, we followed practices and clinicians centrally.

### Encounter SDM tools and the ShareEBM Toolkit

The effective encounter SDM tools targeted by this trial and developed by the Mayo Clinic KER Unit were designed to improve conversations between clinicians and patients regarding medication management for patients with chronic conditions (<u>carethatfits.org/tools</u>). They included Statin Choice (statin use for the primary prevention of cardiovascular disease),^10,10^ Osteoporosis Choice (bisphosphonate use in at-risk women to prevent bone fragility fractures),^12,14^ Diabetes Choice (glucose-lowering medication use in patients with type 2 diabetes)^11,16^ and Depression Choice (antidepressant use in patients with depression).^14^

In close collaboration with intended end-users (patients, clinicians and staff) and other stakeholders (local quality improvement leaders, administrators and executives), we developed a web-based repository, the ShareEBM Toolkit. This toolkit included activities and tactics facilitating the active implementation and maintenance of the targeted encounter SDM tools in practices (**Table 1**). We followed the Replicating Effective Frameworks^24^ and worked iteratively with our end-users and stakeholders, with and without previous experience of SDM or SDM tool usage, from four primary care practices across Minnesota over the course of a one-year period. In the end, these activities and tactics focused on four main categories targeting (1) the clinical encounter (e.g., SDM tools, patient leaflets), (2) clinicians (e.g., online training, guides, tutorials), (3) practices (e.g., journal clubs, presentations), and (4) components to engage leadership (e.g., presentations, performance measures).

**TABLE 1.** ShareEBM toolkit list of activities and tactics.

| SDM encounter tools | TOOLS & RESOURCES | LEADERSHIP | PLANNING & ASSESSING |
| --- | --- | --- | --- |
| **Cardiovascular Primary Prevention Choice (Statin Choice)**<br>• Link to interactive tool<br>• Link to printable DAs<br>• Demos & tutorials<br>• Link to evidence<br>**Depression Med Choice**<br>• Link to printable tool<br>• Storyboard and demos<br>• Take-home brochure<br>• Link to evidence<br>**Diabetes Med Choice**<br>• Link to interactive tool<br>• Link to printable tools<br>• Storyboard & demos<br>• Take-home brochure<br>• Link to evidence<br>**Bone Health Choice**<br>• Frax Score information<br>• Link to interactive tool<br>• Link to printable tools<br>• Demo<br>• Link to evidence | **Journal Clubs Material**<br>• Cardiovascular Prevention Choice<br>• Depression Medication Choice<br>• Diabetes Medication Choice<br>• Bone Health Choice<br>**Video Presentations**<br>• Narrated PowerPoints<br>• 4-min SDM video<br>• 4-min SDM interview<br>**Testimonials**<br>• Clinicians<br>• Patients<br>**EHR templates**<br>• Cardiovascular Prevention Choice<br>• Depression Medication Choice<br>• Diabetes Medication Choice<br>• Bone Health Choice<br>**Related Publications**<br>• Bibliography<br>• References | **Video Presentations**<br>• Narrated PowerPoints<br>**What's-In-It-For-Me**<br>• Briefs<br>**Testimonials**<br>• Clinicians<br>• Patients<br>**Related Publications**<br>• Bibliography<br>• References | **Processes**<br>• Process mapping<br>• Considerations<br>• Worksheets<br>• Value stream mapping<br>• Culture mapping<br>**Measures**<br>• Inventory<br>• Patient satisfaction<br>• Surveys |

DA= Decision aid, SDM = Shared decision making, EHR = Electronic health record

### Intervention (active implementation)

We worked closely with pre-identified practice champions (up to two per practice) and key staff at each practice to actively integrate and promote the use of encounter SDM tools, tailoring activities and tactics from the toolkit to the needs of each practice assigned to this arm. Led by ICSI, we held recurrent individual touchpoints with practices and collective touchpoints with all practice champions to provide updates and share experiences amongst them.

### Control (Passive Dissemination)

We met once with practices assigned to this arm to review study goals and procedures, and present the ShareEBM toolkit. Each practice champions (up to two per practice) was then responsible for the implementation of the encounter SDM tools in their environment, for which they could access the toolkit activities and tactics ad libitum. No further help from the study team was provided.

### Conceptual Frameworks

We supported the evaluation of the impact of the active implementation and passive dissemination strategies using the RE-AIM framework, developed specifically to evaluate how interventions are implemented in a real-world setting.^25^ Dimensions of the RE-AIM include: Reach (how broadly is this intervention used within the practices), Effectiveness (what is the impact of the intervention on outcomes), Adoption (can this be adopted by new groups with ease and minimal modifications), Implementation (what are the special issues and barriers in implementation), and Maintenance (can the intervention be maintained and will the impact continue).^25^

We combined the use of RE-AIM with the Normalization Process Theory (NPT), a sociological theory and whole-system approach to understanding how complex interventions, such as SDM tools, become embedded in practice.^26–28^ NPT focuses on the work individuals or teams do, rather than their attitudes or beliefs, to enable an intervention to become embedded in routines, according to a set of four constructs: Coherence (i.e., sense-making), Cognitive participation (i.e., relational work), Collective action (i.e., operational work), and Reflexive monitoring (i.e., appraisal of the work).^26,27^ NPT constructs were mapped into and added depth of understanding of the Adoption, Implementation, and Maintenance dimensions of the RE-AIM framework.

### Data Collection

We collected quantitative and qualitative data at the practice and clinician levels. We surveyed practice leaders, administrative staff, and clinicians to report on practice and clinician characteristics. We conducted onsite ethnographic observations at 6 months (N=5) and semi-structured interviews at 3 (N=10) and 9 months (N=8) with clinicians or key staff from practices assigned to the intervention arm. We held one focus group (clinicians and staff) at each intervention and control practices at the end of the trial period. Lastly, we collected written documents pertaining to implementation activities from all participating practices (e.g., meeting notes, notices, email exchanges with team members). All qualitative activities were conducted by team members with relevant expertise.

### Outcomes

#### Reach

We assessed the proportion of practices and clinicians who agreed to participate in the study. Clinicians reported “missed opportunities”, i.e., patients for which they could have use a decision aid but did not, and we used these reports to estimate the number of potentially eligible patients. We further documented reach and recruitment approaches through the interviews and focus group with clinicians and staff.

#### Effectiveness

Rate of SDM tool usage by clinicians was our primary outcome. Clinicians tracked their use of each study SDM tool, either through their electronic medical record or log sheet located in their clinical office. We calculated the total number of SDM tools used per arm and per clinician as a fraction of all their potentially eligible encounters. We assessed clinicians’ acceptability and satisfaction at baseline, 3, 6 and at 12 months using a 2-item survey. We further documented usage, acceptability, perception of efficacy, and satisfaction through the interviews and focus group with clinicians and staff.

#### Adoption and Implementation

We assessed the proportion of clinicians who used SDM tools as a fraction of all clinicians who agreed to participate in the study. We tracked use of the tactics and activities of the ShareEBM toolkit or any additional implementation activities through meeting minutes, field notes, and web tracking. We assessed ease of adoption, barriers and facilitators to implementation through ethnographic observations, field notes, interviews, and focus groups with clinicians and staff.

#### Maintenance

We documented the variation of SDM tool use by clinicians over the 12-month period and assessed, at the end of the trial, clinicians and staff perspectives on the level of integration of the SDM tools in practice and their continued use over time. Clinicians completed, at baseline and at 12 months, the Normalization Process Assessment (NPA), a 16-item, 4-domains (coherence, cognitive participation, collective action, reflexive monitoring) questionnaire designed to understand the work that individuals and teams do to enable an intervention to become normalized.^28^

### Sample size

We estimated, for our primary outcome, a conservative 5% rate of use of decision aids within practices assigned to the control arm, raising to 25% within practices assigned to the intervention arm. Assuming (i) a difference in SDM tool use of ≥20% between arms, (ii) a significance level of 0.05 with a two-sided t-test, (iii) a modest correlation of outcomes across clinicians and practices (intra-cluster correlation coefficient [ICC] of 0.05), and (iv) a design effect factor of [1+ (n−1) • ICC] where n is the number of patients per cluster, we estimated the need to recruit 12 practices (30 patients per clinic for a total of 360 patients) to reach 90% power.^29^

### Analysis Plan

We analyzed results according to the intention-to-treat principle.^30^ We report baseline characteristics for practices and clinicians with means and standard deviations for continuous values and counts and frequencies for categorical values. Proportions were compared between group using two-sample t-tests. Clinicians’ satisfaction was modeled using random effect models, clustering for clinic and accounting for the repeated measures while controlling clinician and practice characteristics.

We conducted qualitative analyses by coding verbatim transcripts of clinician interviews and focus groups using a general inductive approach in keeping with RE-AIM and NPT. Analyses were conducted by a team of experienced analysts using the specialized software NVivo 10 (QRS International). We triangulated quantitative and qualitative data for maximum information regarding the extent of adoption and implementation of the targeted decision aids as facilitated by the ShareEBM toolkit.

## RESULTS

We summarized characteristics of practices and clinicians in **Table 2**. There were no significant differences in practice or clinician characteristics across arms.

**Table 2.** Practice and Clinician Characteristics.

|  | PASSIVE DISSEMINATION | ACTIVE IMPLEMENTATION |
| --- | --- | --- |
| PRACTICES, N | N = 6 | N = 5 |
| Location, N (%) |  |  |
| Inner city/ Urban | 2 (33) | 0 (0) |
| Suburban | 3 (50) | 4 (80) |
| Rural | 1 (17) | 1 (20) |
| % Payment Method, median (IQR) |  |  |
| Private health insurances | 23 (5-78) | 74 (37-85) |
| Medicare | 9 (9-35) | 15 (6-23) |
| Medicaid/ Government | 10 (10-11) | 10 (6-35) |
| Others | 1 (1-2) | 1 (0-2) |
| Missing practice(s), N | 1 | 1 |
| Patient Ethnicity, median (IQR) |  |  |
| White | 91 (90-95) | 86 (55-93) |
| Black | 2 (2-4) | 6 (1-20) |
| Others | 3 (0-8) | 3 (0-4) |
| Missing practice(s), N | 1 | 1 |
| Prior experience with SDM tools, N (%) |  |  |
| Participant in a prior KER Unit trial | 1 (17) | 1 (20) |
| Prior use of SDM tools | 1 (17) | 1 (20) |
| CLINICIANS, N | N = 33 | N = 29 |
| Female, N (%) | 27 (82) | 16 (55) |
| Age, Mean (SD) | 48 (10) | 44 (8) |
| Missing, N (%) | 2 (6) | 3 (10) |
| Type of Clinicians |  |  |
| MD/DO | 19 (58) | 19 (66) |
| Other (e.g., advance practitioners, nurses) | 14 (42) | 10 (34) |
| Years in Practice, Mean (SD) | 15(11) | 13 (9) |
| Missing, N (%) | 2 (6) | 2 (7) |
| % in Direct Patient Time, Mean (SD) | 89 (16) | 82 (22) |
| Missing, N (%) | 0 (0) | 2 (7) |
| Estimated Overall Patient Encounters per Month, Mean (SD) | 104 (68) | 100 (65) |
| Missing, N (%) | 5 (15) | 5 (17) |
| Depression encounters | 29 (22) | 24 (19) |
| Missing, N (%) | 5 (15) | 3 (10) |
| Osteoporosis encounters | 9 (10) | 7 (10) |
| Missing, N (%) | 4 (12) | 4 (14) |
| Cardiovascular prevention encounters | 39 (32) | 37 (35) |
| Missing, N (%) | 5 (15) | 3 (10) |
| Diabetes encounters | 30 (23) | 40 (49) |
| Missing, N (%) | 4 (12) | 3 (10) |

### Reach

We approached 45 practices before reaching our targeted 12 (27% reach) practices (**Figure 1**). Twelve practices (27%) did not return our calls or replied to our invitation, 7 (16%) reported being too busy to participate in a research study and 14 (31%) did not provide a reason. One practice from the active implementation arm withdrew after consent but before the start of recruitment period, citing technological challenges and lack of relevance of the selected SDM tool topics for their population (the clinic cared for young migrant families) as reason to withdraw. Four practices (two in each arm) had either participated in a previous KER Unit trial of SDM tools or had experience using other types of decision aids. In the end, we included 11 practices associated with 9 different health systems, within Minnesota and Wisconsin from October 2014 through November 2015.

**Figure 1.**
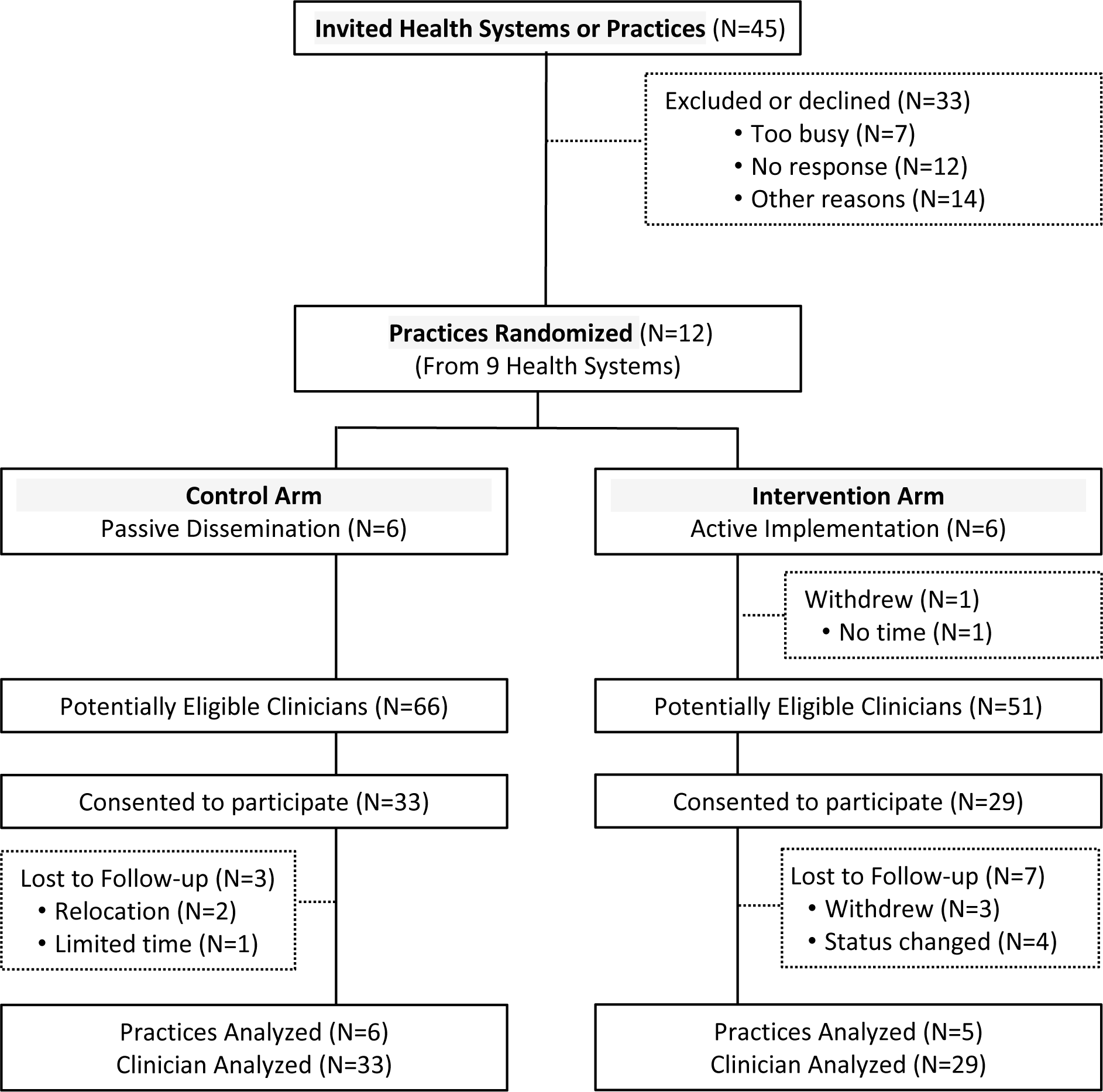
Participants Flow Diagram.

From these participating practices, 62 clinicians (53% of all eligible clinicians) agreed to participate in the study (median [range], 5 [2-11] per practice). There was no difference in the attrition of clinicians or completeness of the data across arms. While we asked practice leaders to invite all eligible clinicians, we were not able to document if all clinicians received an invitation or what were the reasons for declining participation. Furthermore, one practice reported having restricted the number of clinicians allowed to participate in the study, further limiting our capacity to assess with precision clinicians’ reach.

Patients’ reach was under clinicians’ responsibility as they were given discretion of when to use the SDM tools. Over the 12-month period, clinicians reported that they could have used a tool with 41 222 (passive dissemination) and 38 967 (active implementation) patients seen in their offices. Our choice to make encounter eligibility criteria minimal (i.e., any encounter in which a medication management decision is likely to occur) to facilitate recruitment had however the unintended consequence, revealed in interviews with clinicians, of not providing specific criteria that could have resulted in proactive identification – electronically or by a member of the healthcare team – of encounters eligible for SDM tool use.

### Effectiveness

Clinicians used a total of 680 SDM tools in the passive dissemination arm compared to 621 in the active implementation arm, with no difference in the average rate of use across arms (**Table 3**). Statin Choice was the most commonly used SDM tool (75% of all tools used) in both arms. Satisfaction and intent to recommend SDM tools to others increased over time across both arms.

**Table 3:**
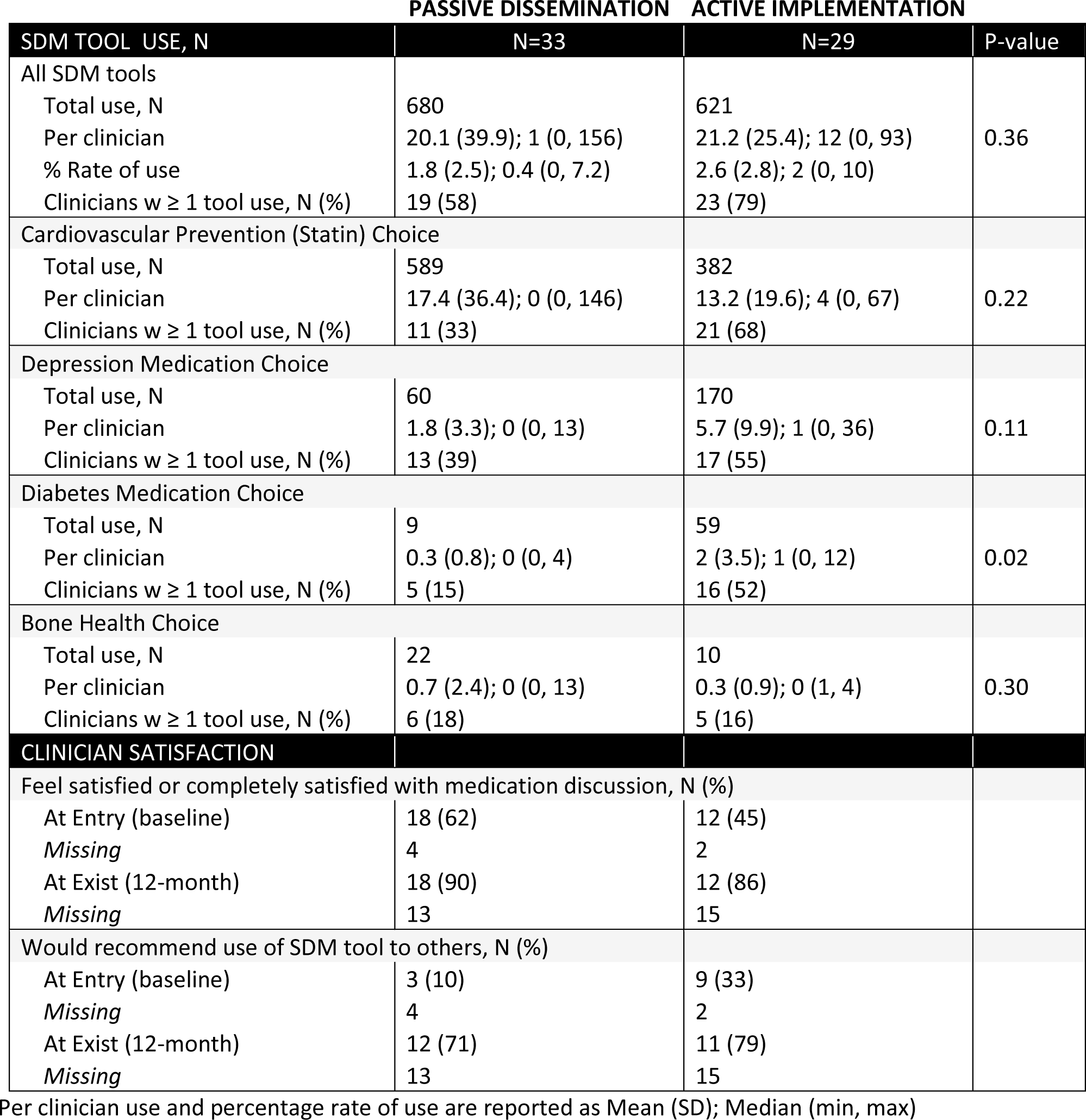
Clinicians Use of SDM tools and Satisfaction.

### Adoption, Implementation, and Maintenance

Adoption of SDM tools by practices and clinicians was uneven across arms. Almost all SDM tool use (95%) took place in two practices (n=503 and 146) in the passive dissemination arm (**Table 3**). Usage was somewhat balanced across practices in the active implementation arm with the most active practice accounting for 49% (n=306) of SDM tool use. The two leading practices, although in different arms, were affiliated with the same health system. Fewer clinicians in the passive dissemination arm used SDM tools compared to those in the active implementation one, with one outlier clinician (n=156) driving total use (**Figure 2**). Decision aid usage varied over time in both arms, with no discernable patterns (**Figure 3**).

**Figure 2.**
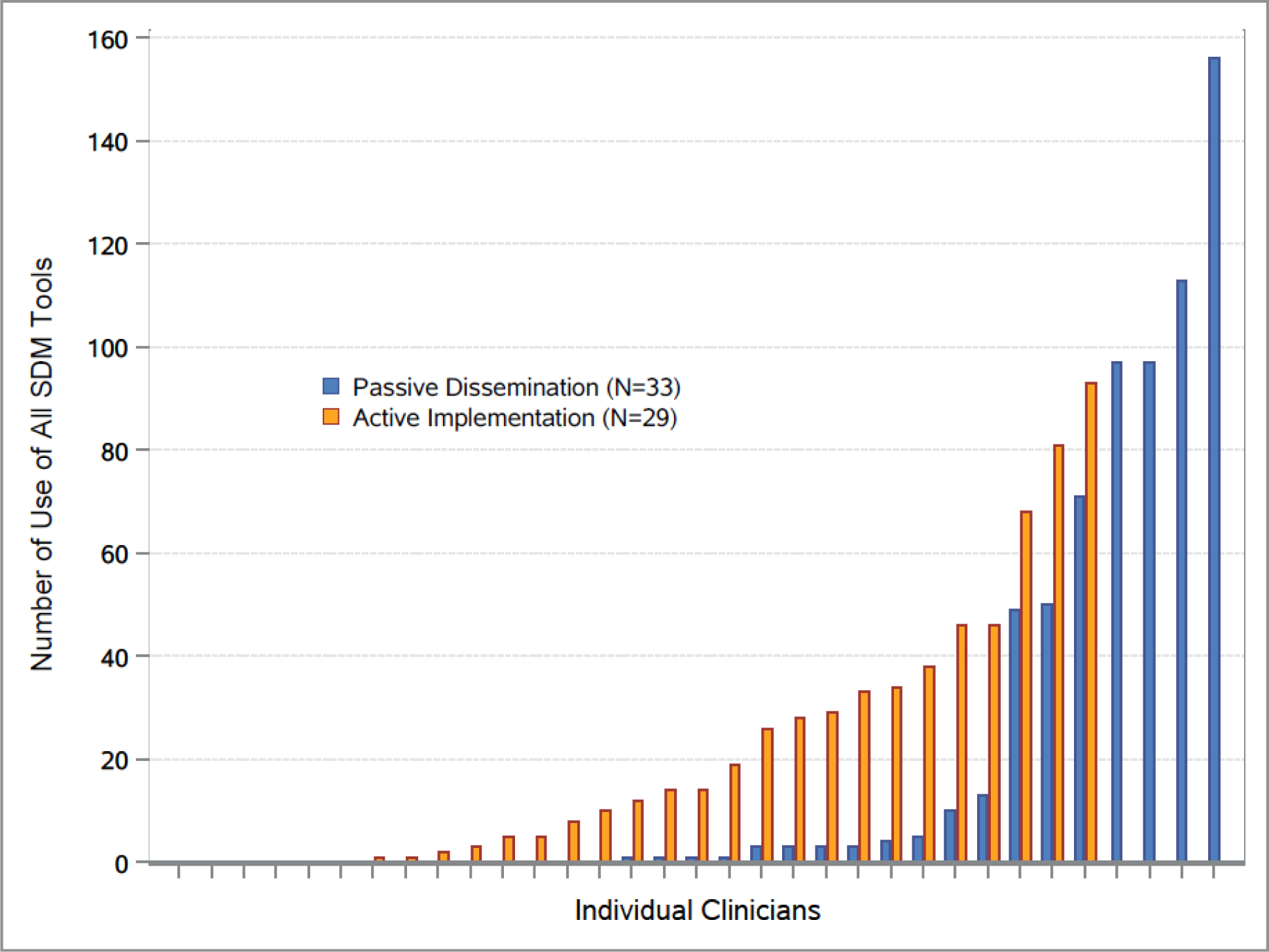
SDM tool usage per clinician.

**Figure 3.**
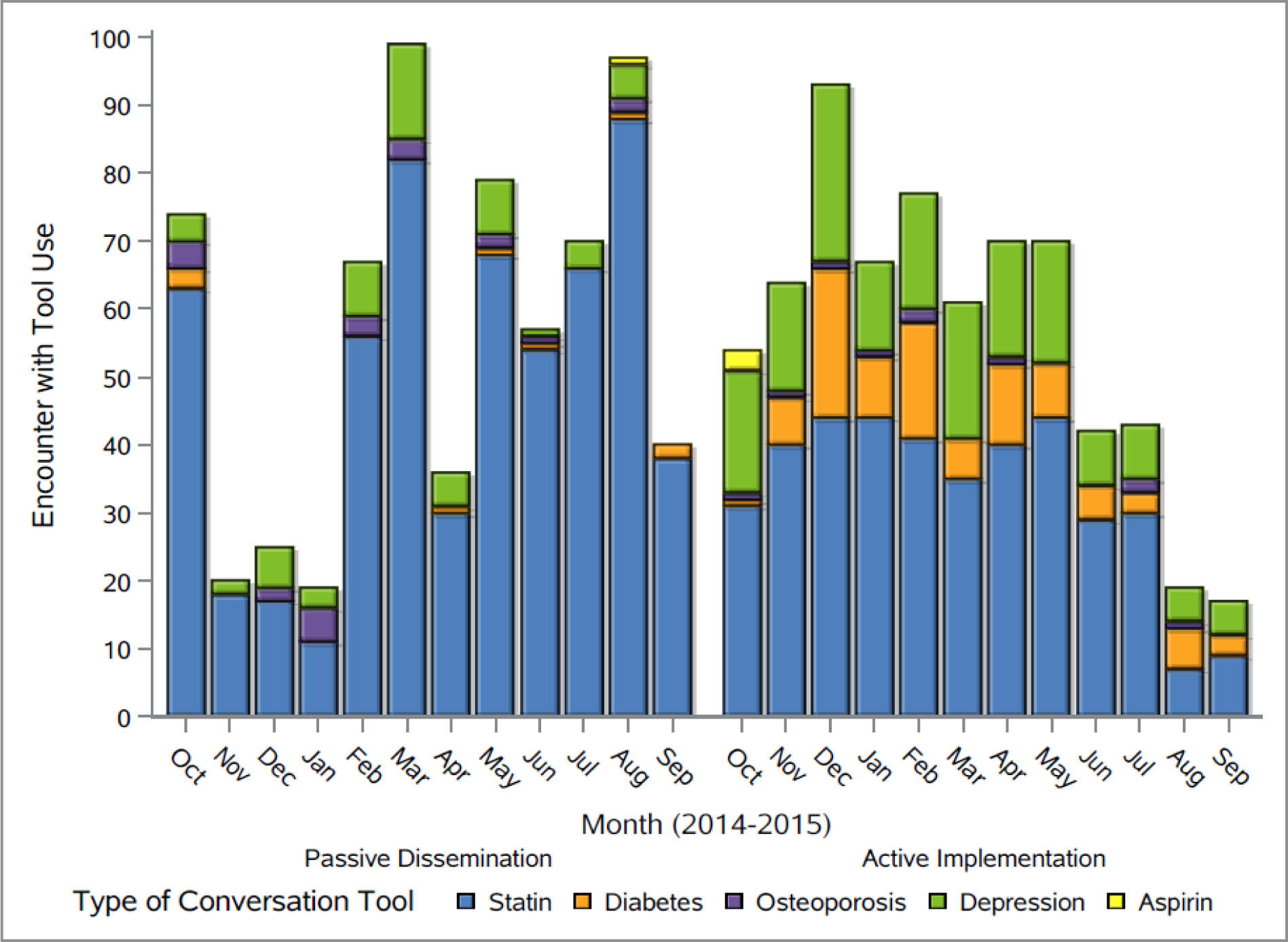
SDM tools usage over time across arms.

According to the website login information, the use of the ShareEBM toolkit activities and tactics was limited to accessing the SDM tools themselves and journal club material across arms. None of the other activities or tactics from the toolkit were requested directly or addressed during touchpoints or focus groups.

Clinicians’ initial assessment of the potential of SDM tool integration and normalization into their practices was high across all domains, with reflexive monitoring (i.e., evaluating the use of the SDM tools in daily practice and monitoring its value) scoring somewhat lower across arms (**Figure 4**). Post-intervention, clinicians in the passive dissemination arm offered lower-than-baseline ratings to all domains of normalization, except from the coherence domain (i.e., how using SDM tools made sense in their practice) which remained unchanged. In contrast, except from the reflexive monitoring domain, clinicians in the active implementation arm offered higher-than-baseline rating for all domains.

**Figure 4.**
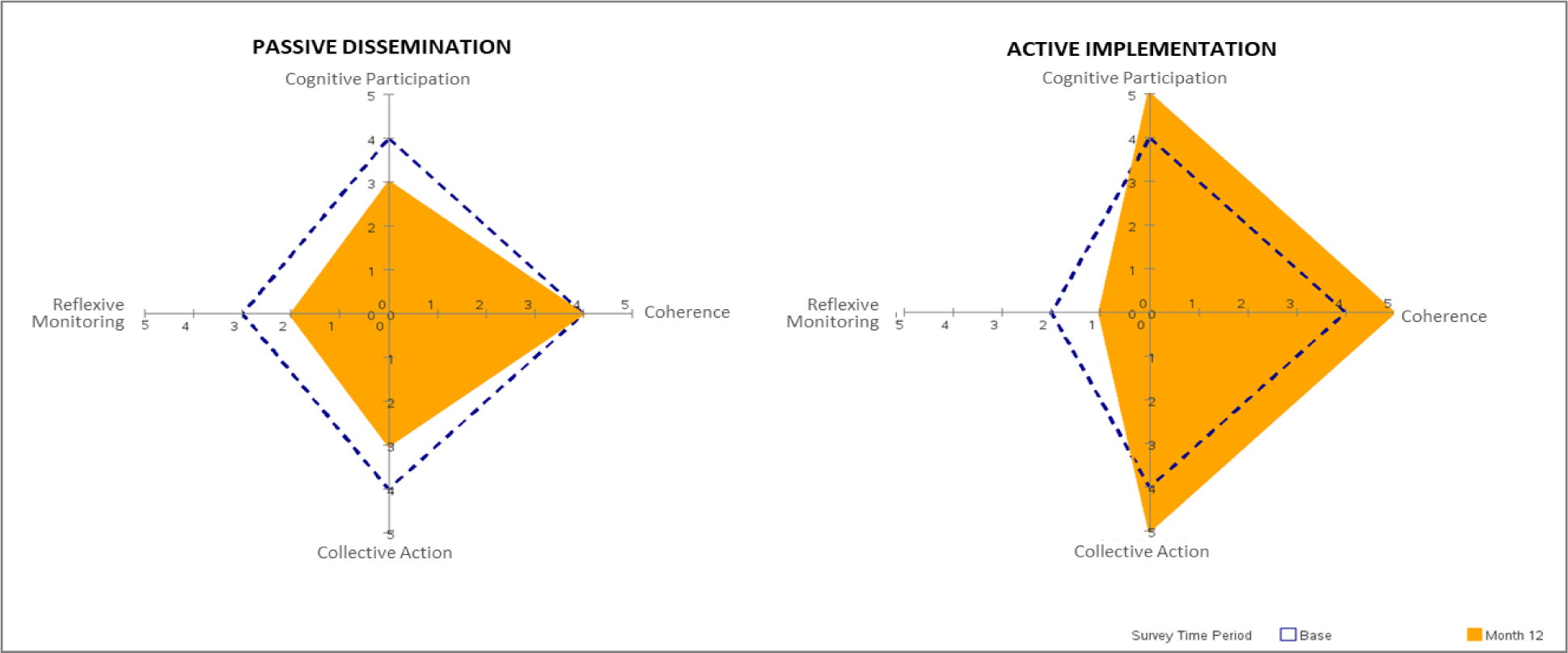
Normalization process assessment (NPA) pre and post SDM tools usage.

Interviews, focus groups, and field notes revealed that adoption and implementation varied considerably within and across practices. Four themes emerged as central to the adoption and implementation of SDM tools: clinicians’ general views of SDM tools, the impact they perceived these tools could have on patient care, how they used the tools, and how they incorporated SDM tools into their workflow (**Table 4**).

**Table 4.**
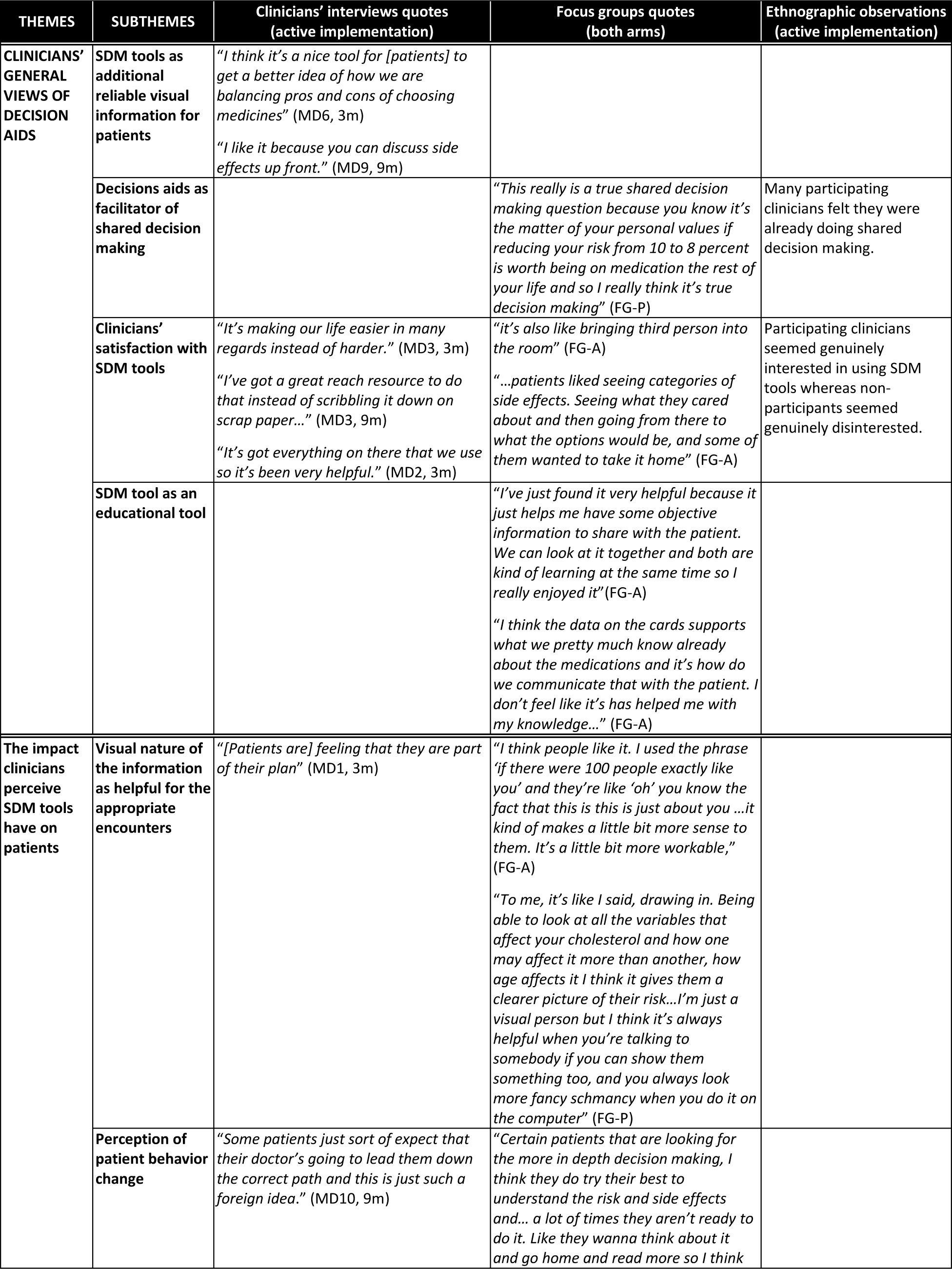
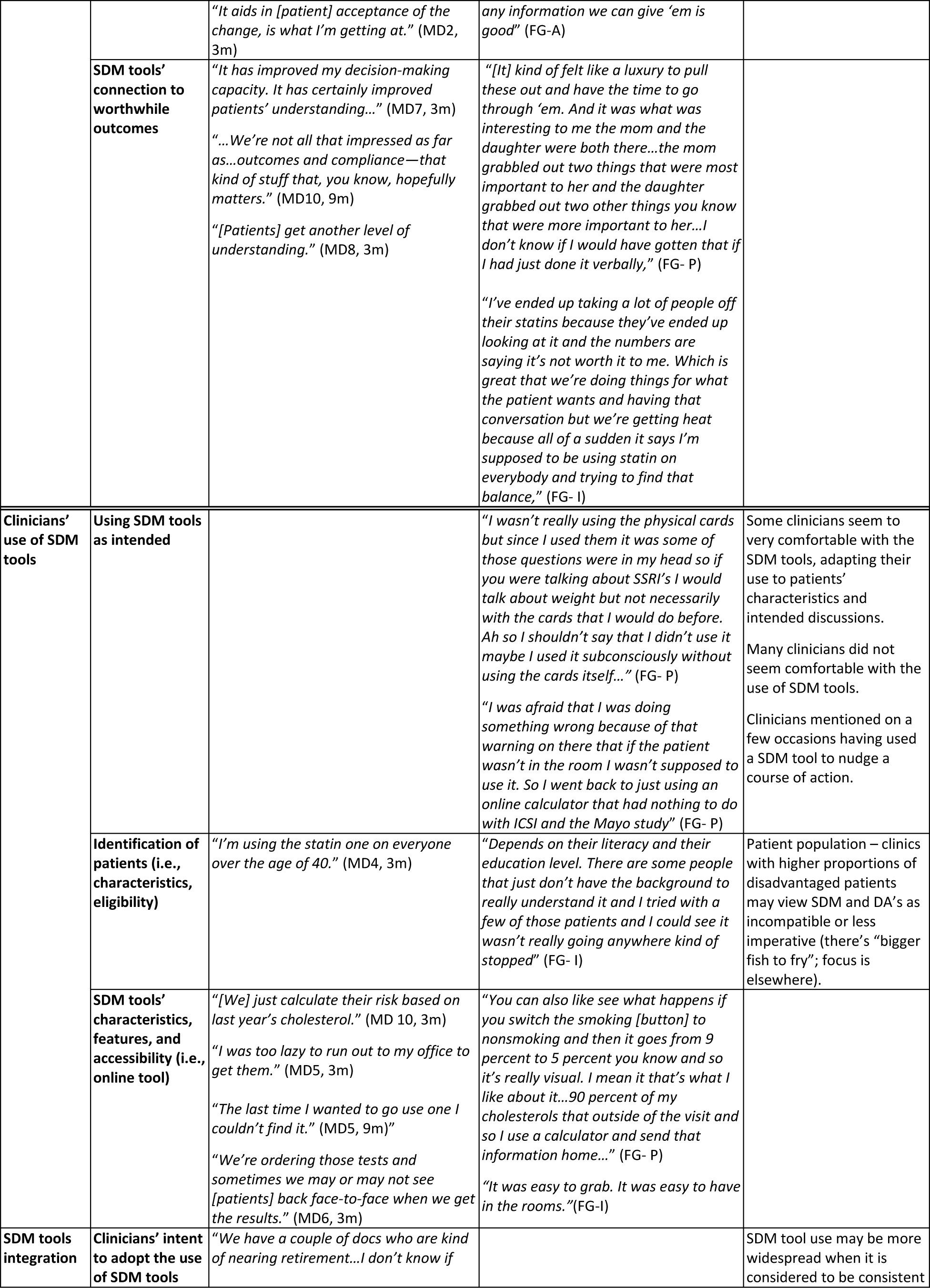
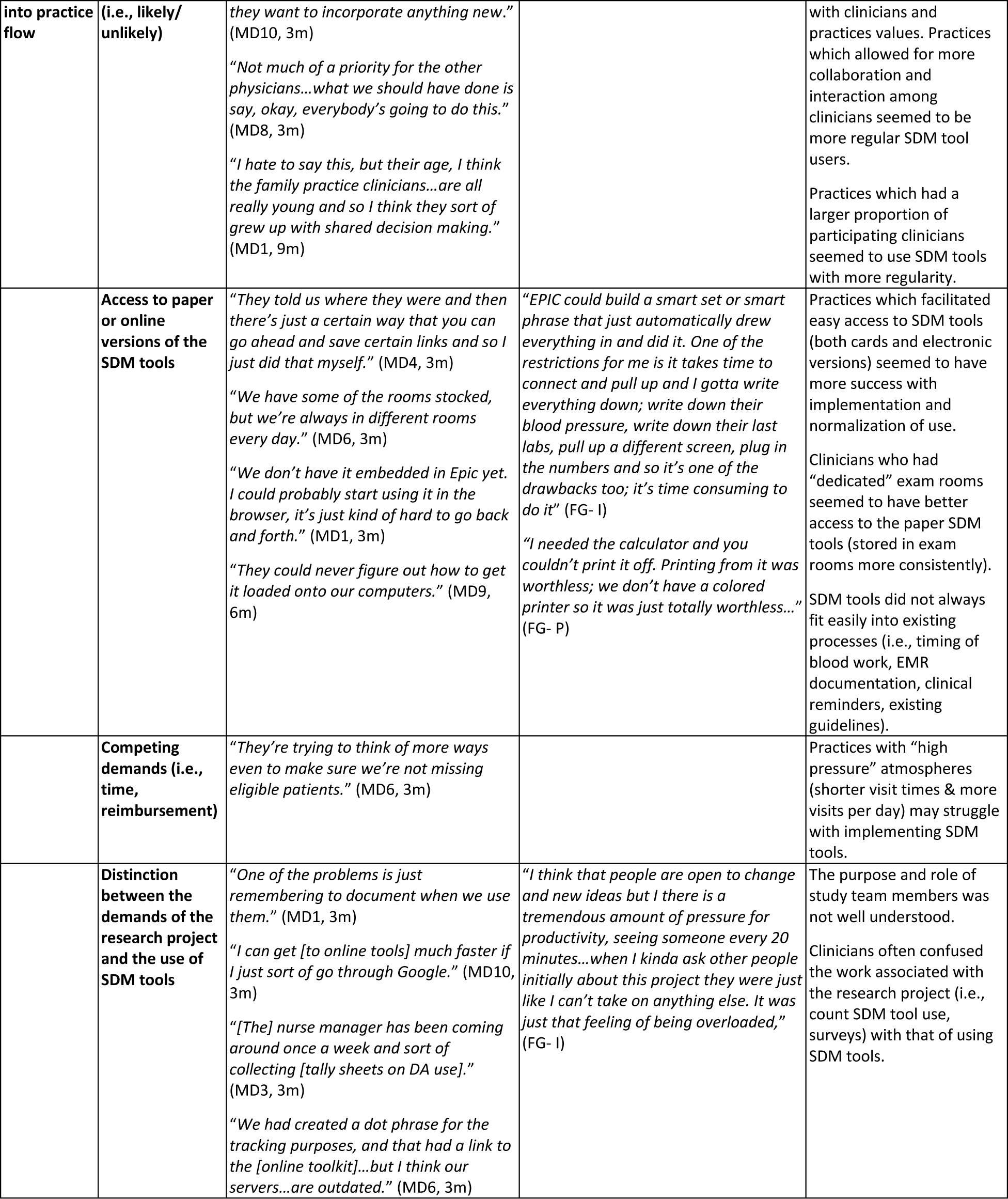
Themes and quotes associated with the adoption and implementation of SDM tools.

Overall, participating clinicians felt they were already engaging their patients in SDM. They thought SDM tools were well-designed, provided reliable visual information, and facilitated SDM. However, not all clinicians were convinced that using SDM tools would lead to worthwhile outcomes, thus judging their use in practice not worth the time. Accessing SDM tools, whether the paper or online versions, seemed to be the main driver of use and key determinant of normalization into the workflow. While some clinicians were very comfortable with the use of SDM tools, adapting their use to patient characteristics, discussions about how they used the tools with their patients suggest that more extensive training and role modeling may have been warranted.

There was a general feeling from all participating practices and clinicians that there were likely adopters who will do everything and work around barriers to use SDM tools and unlikely adopters who will not use them “no matter what”. Likelihood of adoption was further tainted by demands perceived as competing (e.g., reimbursement), time constraints, and the perceived ability of their patient population to take part in SDM. The likelihood of long-term use appears connected to the ability to integrate SDM tools into the electronic and practice workflows; there was general agreement from participating practices that this is something worth doing (i.e., embedding SDM within the practice culture and processes) (**Table 4**).

To some extent, the research study and intervention itself (ShareEBM Toolkit) were limiting factors. While a few practices took advantage of this project to embark on a practice-wide implementation of SDM tools, other practices saw this as “another” research project with leaders minimizing engagement and limiting clinician participation (**Table 4**). Some clinicians and practices did not make the distinction between the work associated with the use of the SDM tools, which would remain after the study was over, and the work associated with the trial itself (e.g., filling surveys, tracking usage), which may explain the lower score observed for the reflexive monitoring domain of the NPA. The setup of the intervention (access to the toolkit necessitated a login process for research tracking purpose) created confusion about the free access and use of the SDM encounter tools.

## DISCUSSION

### Main findings

In this pragmatic randomized trial, an SDM implementation toolkit whether actively or passively implemented, did not significantly contribute to the uptake and use of SDM encounter tools by clinicians or practices. Likelihood of long-term use of encounter SDM tools appears connected to the ability to integrate the tools into electronic and practice workflows, provide feedback on fidelity and quality of their use during encounters, and overcoming clinicians’ reluctance to use SDM tools.

### Comparison with other studies

Our results are comparable to previous implementation initiatives.^19–21,31^ As others, we found that adoption requires organizational support, local ownership and leadership, and above all, a favorable attitude and openness toward SDM.^32,33^ Sparse access and use of the activities and tactics, which targeted clinicians, suggest that a larger focus on the practice cultural may be warranted.

Clinicians’ perceptions that they are already doing SDM and their view that using SDM tools does not add enough value to justify the additional work of accessing and using them in practice are findings also consistent with previous work.^18,20,23,32,34^ Furthermore, in our study, clinicians insisted on the importance of just-in-time access to the SDM tools, a characteristic highlighted in studies for more than a decade now, with few reporting on successfully implementing easy access and use within electronic and practice workflows.^21,35^ ^36^

### Limitations

Our rigorous design did not fully protect our study from the intrusion of different sources of error. The cluster design did not protect the trial from patient or clinician selection bias. Our findings may not be applicable to other primary care settings or contemporary practices. Where possible, we sought to overcome these limitations through triangulation of methods and seeking self-reported data.

## CONCLUSION

We intended to develop an implementation toolkit, ShareEBM, that would contribute to improve care quality by supporting primary care practices on their implementation of encounter SDM tools in the care of patients with chronic conditions. Our findings, supported by a growing literature on encounter SDM tools, provided insights on the complexity of implementing SDM tools at the point of care and the importance of operational and cultural aspects in the success of these efforts.

## Data Availability

The SDM encounter tools can be freely accessed on the Mayo Clinic Shared Decision Making National Resource Center website (http://carethatfits.org). The ShareEBM activities and tactics can be requested from the authors. De-identified data can be requested and provided to investigators upon alignment with and approval from Mayo Clinic Ethics Committee and legal authorities.

## DECLARATIONS

### Ethics approval and consent to participate

The Mayo Clinic Institutional Review Board approved study procedures (14-002055).

### Authors’ contributions

AL, MEB, NDS, JS, and VMM contributed to the study concept and design and sought funding. SD, JS and JK contributed to the data collection. JWI and MEB conducted the statistical analysis. JE contributed to the qualitative analysis. AL drafted the first version of the manuscript. All provided critical revision of the manuscript. AL and VMM take responsibility for the integrity of the data and the accuracy of the data analysis and reporting.

## ACKOWLEDGEMENT

We would like to acknowledge Juliette Liesinger, Megan Morris, Ashok Kumbama, and Cally Vinz for their support on this project.

